# Genomic Sequencing for Newborn Screening: Results of the NC NEXUS Project

**DOI:** 10.1101/2020.02.26.20024679

**Authors:** Tamara S. Roman, Stephanie B. Crowley, Myra I. Roche, Ann Katherine M. Foreman, Julianne M. O’Daniel, Bryce A. Seifert, Kristy Lee, Alicia Brandt, Chelsea Gustafson, Daniela M. DeCristo, Natasha T. Strande, Lori Ramkissoon, Laura V. Milko, Phillips Owen, Sayanty Roy, Mai Xiong, Ryan S. Paquin, Rita M. Butterfield, Megan A. Lewis, Katherine J. Souris, Donald B. Bailey, Christine Rini, Jessica K. Booker, Bradford C. Powell, Karen E. Weck, Cynthia M. Powell, Jonathan S. Berg

## Abstract

Newborn screening (NBS) was established as a public health program in the 1960’s and is crucial for facilitating detection of certain medical conditions in which early intervention can prevent serious, life-threatening health problems. Genomic sequencing can potentially expand the screening for rare hereditary disorders, but many questions surround its possible use for this purpose. We examined the use of exome sequencing (ES) for NBS in the <u>N</u>orth <u>C</u>arolina <u>N</u>ewborn <u>Ex</u>ome Sequencing for <u>U</u>niversal <u>S</u>creening (NC NEXUS) project, comparing the yield from ES used in a screening versus a diagnostic context. We enrolled healthy newborns and children with metabolic diseases or hearing loss (106 participants total). ES confirmed the participant’s underlying diagnosis in 15 out of 17 (88%) children with metabolic disorders, and in 5 out of 28 (∼18%) children with hearing loss.

We discovered actionable findings in 4 participants that would not have been detected by standard NBS. A subset of parents was eligible to receive additional information for their child about childhood-onset conditions with low or no clinical actionability, clinically actionable adult-onset conditions, and carrier status for autosomal recessive conditions. We found pathogenic variants associated with hereditary breast and/or ovarian cancer in 2 children, a likely pathogenic variant in the gene associated with Lowe syndrome in one child, and an average of 1.8 reportable variants per child for carrier results. These results highlight the benefits and limitations of using genomic sequencing for NBS and the challenges of using such technology in future precision medicine approaches.

## Introduction

Newborn screening was established in the early 1960’s, beginning with phenylketonuria^1^ (PKU) (MIM: 261600), for which early intervention can prevent permanent intellectual disability. In the 1990’s tandem mass spectrometry was employed to efficiently screen neonates for numerous metabolites, which resulted in an increase in the number of genetic conditions screened^2–4^. As of July 2018, The American College of Medical Genetics and Genomics and the United States federal Advisory Committee on Heritable Disorders in Newborns and Children have identified 35 core conditions and 26 secondary conditions to be included on the recommended uniform screening panel (RUSP, recommended by the Secretary of the Department of Health and Human Services), including specific disorders of organic acids, fatty acid oxidation, amino acids, as well as endocrine and hemoglobin disorders^5–7^. Most infants in the United States also receive point of care newborn screening for hearing and critical congenital heart disease.

Advances in genomic sequencing technologies (next-generation sequencing, or NGS), such as exome and genome sequencing (ES or GS, respectively), have emerged as powerful tools for identifying an individual’s genomic changes at the DNA base level^8^. These technologies have the potential to dramatically increase the ability to screen newborns for rare hereditary disorders that would not be detected using traditional screening methods. Given this potential, the National Institute of Child Health and Human Development (NICHD) and the National Human Genome Research Institute (NHGRI) sponsored a consortium of studies, the Newborn Sequencing in Genomic Medicine and Public Health (NSIGHT) consortium, to investigate the use of this technology in newborns^9–12^. One of these studies recently reported on the use of NGS in healthy newborns and reported additional findings that would have been missed by existing newborn screening modalities^10^. Thus, for some cases, genomic sequencing may clarify the underlying genetic cause of an individual’s disease and directly impact a patient’s diagnosis, intervention and/or treatment. In addition, genomic sequencing may also be useful for screening asymptomatic individuals.

In the North Carolina Newborn Exome Sequencing for Universal Screening (NC NEXUS) study, we explored the possible use of ES in the context of newborn screening and studied the responses of parents to this new technology. The study design^13^ included two cohorts of infants and children: 1) healthy newborns whose parents were approached for participation in the study prenatally; and 2) infants and children (< 5 years of age) already clinically diagnosed with conditions detected with current newborn screening methods (inborn errors of metabolism and hearing loss).

In the current manuscript, we report the molecular analysis results from the cohort of 106 children who were enrolled in the study. Molecular analysts examined ES data using a next-generation sequencing newborn screen (NGS-NBS) panel of 466 genes^14^, while being blinded to the identity of the participant’s cohort (well-child, metabolic or hearing loss). In our analysis strategy, we defined strict criteria for assessing the screen as “positive,” which allowed us to examine the sensitivity of ES when used as a screening test to detect a genetic alteration in the context of a theoretical newborn screen. For those participants in the metabolic cohort or the hearing loss cohort, we then performed a second, indication-based, diagnostic analysis assessing the ES results for additional genes that were likely associated with the participant’s phenotype. In addition, parents were randomized to one of two study arms. In one study arm, parents could elect to learn additional findings from their child’s ES. The results of the initial NGS-NBS screening, indication-based analyses and additional genomic findings are presented in this work.

## Methods

### Recruitment and sample collection

The study protocol was approved by the University of North Carolina Institutional Review Board and all participants were consented to the study by a certified genetic counselor. The NC NEXUS randomized controlled trial protocol and study design were previously described^13^. As part of that protocol, 17 children previously diagnosed with inborn errors of metabolism and 28 children previously diagnosed with hearing loss were recruited into the study. Additionally, we recruited parents in the prenatal period (pregnancies of 18 weeks or longer) in which the fetus did not have a positive or pending chromosomal abnormality or congenital malformation diagnostic test result. The mothers subsequently gave birth to 61 infants; these babies were classified into the well-child cohort. Participant demographic information is reported in Supplemental Table 1.

**Table 1:**
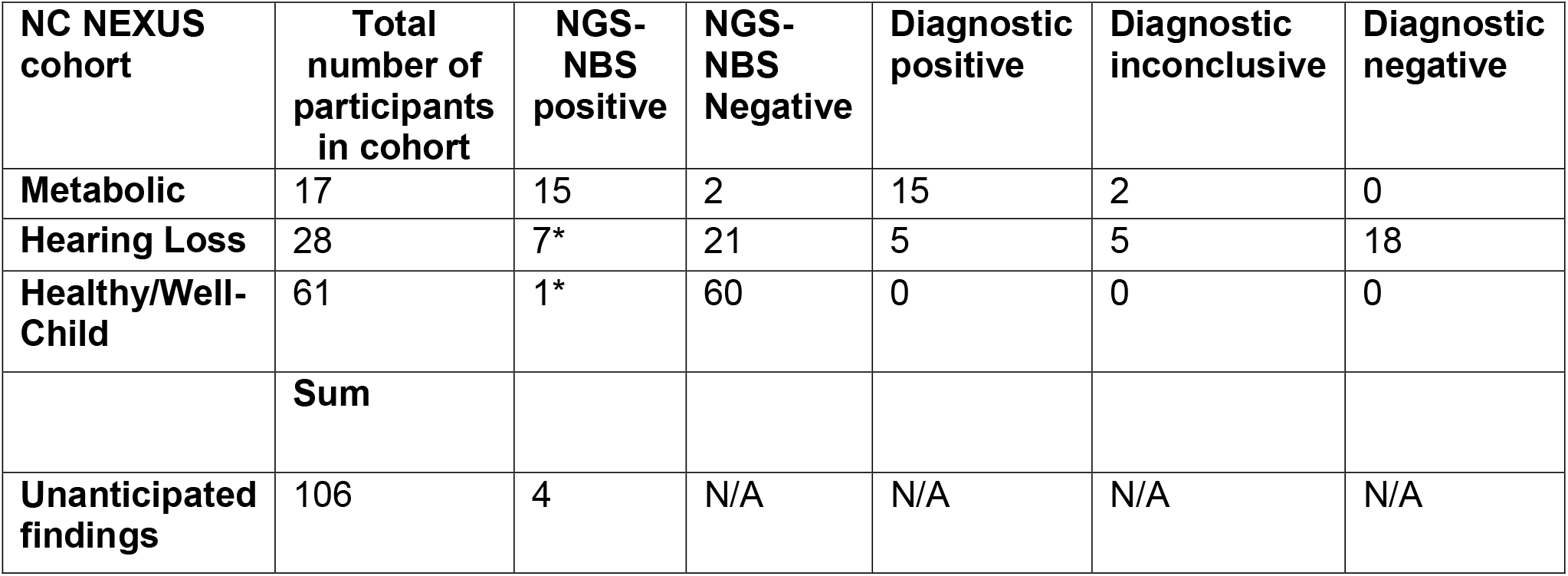
Summary of NGS-NBS and diagnostic findings in the NC NEXUS study. NGS-NBS positive indicates an abnormal, “positive screen,” defined by observing likely pathogenic and/or pathogenic variants in 466 genes associated with pediatric conditions with high medical actionability. NGS-NBS negative indicates a “negative screen,” defined by the absence of likely pathogenic or pathogenic variants found in these genes. Diagnostic positive indicates the presence of likely pathogenic or pathogenic variants found in gene(s) on the metabolic or hearing loss diagnostic list that are consistent with the participant’s disorder. Diagnostic inconclusive indicates an inconclusive result, which was defined as a single heterozygous variant found in a gene associated with an autosomal recessive condition and/or the presence of variants of uncertain significance in genes from the diagnostic list. Numbers with asterisks include two participants in the hearing loss cohort with a positive NGS-NBS due to likely pathogenic variants in *DSC2* or *F11*, and include one participant in the healthy/well-child cohort that was determined to have a positive NGS-NBS screen due to a pathogenic variant in *LDLR* associated with familial hypercholesterolemia. NGS-NBS= next-generation sequencing newborn screen.

| NC NEXUS cohort | Total number of participants in cohort | NGS-NBS positive | NGS-NBS Negative | Diagnostic positive | Diagnostic inconclusive | Diagnostic negative |
| --- | --- | --- | --- | --- | --- | --- |
| Metabolic | 17 | 15 | 2 | 15 | 2 | 0 |
| Hearing Loss | 28 | 7* | 21 | 5 | 5 | 18 |
| Healthy/Well-Child | 61 | 1* | 60 | 0 | 0 | 0 |
|  | Sum |  |  |  |  |  |
| Unanticipated findings | 106 | 4 | N/A | N/A | N/A | N/A |

### Decision aids

As part of the overall study, we previously described a decision aid for parents who would be offered ES for their child^13,15^. This web-based electronic decision aid was developed to enhance parents’ abilities to make decisions about learning genomic information for their child; the aid included both an educational component and a values clarification exercise. The educational component included information about newborn screening, genomic sequencing and the process of using genomic sequencing to identify genetic variants associated with specific diseases. This educational component was followed by a values clarification exercise in which parents classified 5 reasons for and 5 reasons against having genomic sequencing for their child by level of importance^15–17^. After viewing the decision aid, parents met face-to-face with a genetic counselor for additional clarification before being asked to decide whether to consent to ES for their child.

### Age-based semi-quantitative metric

As previously described^14^, we determined actionability of gene-disease pairs using the age-based semi-quantitative metric (ASQM). Conditions were placed into one of four categories: Category 1: pediatric conditions with high medical actionability; Category 2: pediatric conditions with low or no medical actionability; Category 3: adult conditions with high medical actionability; and Category 4: adult conditions with low or no medical actionability. In total, 822 gene-disease pairs were assessed, enriched for those with pediatric onset of disease and suspected actionability. Category 1 (466 gene-disease pairs) included many core RUSP conditions, as well as other disorders with onset in infancy or in childhood that have treatment, monitoring and/or medical management that can potentially improve clinical outcomes; these conditions are comparable to those detected by current NBS and therefore all NC NEXUS study participants were eligible for disclosure of positive findings.

### Randomization for additional findings

Parents were randomized to one of two study arms. One arm was the “Control arm” (1/3 of participants) in which parents learned findings from the NGS-NBS panel and, if applicable, indication-based analysis. The second arm was the “Decision arm” (2/3 of participants) in which parents learned findings from the NGS-NBS (and indication-based analysis if applicable) and were also asked to decide whether or not they wanted to learn about additional types of genomic findings from up to three categories. These three categories included: childhood-onset conditions with low or no clinical actionability, adult-onset conditions with high actionability, and carrier status for autosomal recessive disorders.

### Exome sequencing

The BioSpecimen Processing Facility at the University of North Carolina at Chapel Hill isolated genomic DNA from saliva and oral epithelial cell samples using PureGene chemistry. Exome libraries, including molecular barcoding and exome capture, were prepared using Agilent SureSelect XT kits (Human All Exon V6 probes) according to the manufacturer’s low input guidelines. The University of North Carolina High-Throughput Sequencing Facility performed exome sequencing on an Illumina HiSeq2500 with minimum depth of 40X coverage. Raw sequence reads were mapped to the human genome reference GRCh38.p7 using BWA-MEM version 0.7.12 (arXiv:1303.3997v1 [q-bio.GN]), duplicate reads were marked using Picard MarkDuplicates (version 1.130) and variants were called using freebayes, version 1.1.0 (arXiv preprint arXIv:1207.3907 [q-bio.GN] 2012).

### Bioinformatics and Molecular Analysis

Variants were prioritized based on previous classification as pathogenic (P) or likely pathogenic (LP) in a ClinVar submission^18^, minor allele frequency in a reference database^19^ and predicted effect of the variant on the protein (frameshift, nonsense, canonical splice-site, missense, synonymous, intronic variants). All rare possibly damaging variants in 466 genes in Category 1 were reviewed by a molecular analyst for each of the 106 participants. For the initial NGS-NBS analysis, molecular analysts were blinded to the participant’s cohort (metabolic, hearing loss, or well-child). Variants were classified based on ACMG/AMP interpretation guidelines^20,21^ using collected evidence from population databases, ClinVar^18^ and the primary literature. Pathogenic and/or LP variants that were considered an abnormal, “positive” screen (heterozygous P/LP variants in a gene associated with a dominant condition; two or more P/LP variants in a gene associated with a recessive condition) were flagged for further review by a committee of molecular geneticists and physicians, genetic counselors and researchers. Variants of uncertain significance (VUS) were not returned in the NGS-NBS screen. After completion of the first NGS-NBS analysis, the participant’s cohort was revealed to the molecular analyst. If the participant was part of the metabolic cohort or hearing loss cohort, ES data were further analyzed by filtering for variants in a subset of genes within indication-based “diagnostic lists,” containing genes associated with the relevant phenotypes. For the indication-based diagnostic analyses, P, LP and VUS were reported. All variants were confirmed in a duplicate sample using an orthogonal method performed in the UNC Hospitals Molecular Genetics CLIA-certified laboratory. Copy number variant analysis was not conducted due to restrictions based on the study’s Investigational Device Exemption approved by the FDA^13^.

For participants who decided to learn some or all of the additional findings categories, ES data were analyzed to identify P/LP variants in 22 genes associated with adult-onset actionable conditions, 234 genes associated with childhood-onset nonactionable conditions, and/or 871 genes associated with carrier findings for autosomal recessive conditions.

All parents were seen by a genetic counselor and/or clinical geneticist for return of any positive results in diagnostic, NGS-NBS, childhood-onset medically non-actionable and adult-onset medically actionable categories. Positive results for carrier status were conveyed by a genetic counselor only by phone. Recommendations and options for further testing or evaluation were provided but not part of the study. Parents were given a copy of the CLIA-confirmed result report for their child and were given the option of having the results placed in their child’s electronic health record.

## Results

Molecular analysis was completed for 106 participants: 61 in the healthy/well-child cohort, 17 in the metabolic cohort and 28 in the hearing loss cohort. A total of 43 out of 46 variants detected by ES were confirmed by orthogonal testing in the CLIA-certified laboratory prior to reporting. Additionally, 5 variants that were previously identified by clinical molecular testing were also detected by ES.

### Metabolic cohort

NGS-NBS analysis was deemed to represent an abnormal, “positive screen” result in 15 of 17 participants (88%) in the metabolic cohort (Tables 1 and 2). These individuals had a combination of pathogenic and/or likely pathogenic variants in genes implicated in metabolic conditions, that, when blinded to the participants’ cohort, were determined to be reportable in a screening setting.

**Table 2:** NGS-NBS and diagnostic findings in the NC NEXUS inborn errors of metabolism cohort. The NC NEXUS participant column includes “M” (indicating inborn errors of metabolism cohort) followed by the participant number. Each row in the table represents one NC NEXUS participant. Positive NGS-NBS results indicate a likely pathogenic or pathogenic variant found in the newborn screen. A positive diagnostic result indicates a likely pathogenic or pathogenic variant in a gene on the inborn errors of metabolism diagnostic list. An inconclusive diagnostic result is indicated by any variant of uncertain significance (VUS) finding or a single heterozygous variant found in a gene associated with autosomal recessive inborn errors of metabolism. NGS-NBS= next-generation sequencing newborn screen; AR= autosomal recessive pattern of inheritance; XL= X-linked pattern of inheritance; PKU= phenylketonuria; MCAD deficiency= medium-chain acyl-coA dehydrogenase deficiency; MSUD= maple syrup urine disease.

| NC NEXUS participant | NGS-NBS result | Diagnostic (Dx) result | Gene(s) | Disease association with gene (inheritance) | Variant and (predicted protein change) | Zygosity in NC NEXUS participant | Genomic coordinates (hg38) of variant(s) |
| --- | --- | --- | --- | --- | --- | --- | --- |
| M001 | Positive | positive | *ACADM* | MCAD deficiency (AR) | c.799G>A (p.G267R); c.985A>G (p.K329E) | heterozygous for both variants | chr1:75749509; chr1:75761161 |
| M002 | Positive | positive | *PAH* | PKU (AR) | c.1315+1G>A (p.?); c.782G>A (p.R261Q) | heterozygous for both variants | chr12:102840399; chr12:102852875 |
| M003 | Positive | positive | *PAH*,<br>*OTC* | PKU (AR),<br>OTC deficiency (XL) | *PAH* c.1222C>T (p.R408W) and c.896T>G (p.F299C); *OTC* c.1061T>G (p.F354C) | heterozygous for *PAH* and *OTC* variants | chr12:102840493; chr12:102851703; chrX:38421078 |
| M004 | Positive | positive | *PAH* | PKU (AR) | c.1315+1G>A (p.?); c.284_286delTCA (p.I95del) | heterozygous for both variants | chr12:102840399; chr12:102894805-102894807 |
| M005 | Positive | positive | *PAH* | PKU (AR) | c.194T>C (p.I65T);<br>c.814G>T<br>(p.G272*) | heterozygous<br>for both variants | chr12:102894893;<br>chr12:102852843 |
| M006 | Positive | positive | *ACADM* | MCAD<br>deficiency<br>(AR) | c.985A>G<br>(p.K329E) | homozygous | chr1:75761161 |
| M007 | Positive | positive | *PAH* | PKU (AR) | c.1315+1G>A<br>(p.?); c.842C>T<br>(p.P281L) | heterozygous<br>for both variants | chr12:102840399;<br>chr12:102852815 |
| M008 | Positive | positive | *ACADM* | MCAD<br>deficiency<br>(AR) | c.985A>G<br>(p.K329E) | homozygous | chr1:75761161 |
| M009 | Negative | inconclusive<br>(VUS) | *MLYCD* | Malonyl-coA<br>decarboxylase<br>deficiency<br>(AR) | c.1013T>C<br>(p.L338P) | homozygous | chr16:83915020 |
| M010 | Positive | positive | *PAH* | PKU (AR) | c.117C>G<br>(p.F39L);<br>c.842C>T<br>(p.P281L) | heterozygous<br>for both variants | chr12:102912842;<br>chr12:102852815 |
| M011 | Positive | positive | *PAH* | PKU (AR) | c.194T>C (p.I65T) | homozygous | chr12:102894893 |
| M012 | Positive | positive | *ACADM* | MCAD<br>deficiency<br>(AR) | c.985A>G<br>(p.K329E) | homozygous | chr1:75761161 |
| M013 | Positive | positive | *ACADM* | MCAD<br>deficiency<br>(AR) | c.985A>G<br>(p.K329E) | homozygous | chr1:75761161 |
| M014 | Positive | positive | *SCL22A5* | Renal<br>carnitine<br>transport<br>deficiency<br>(AR) | c.506G>A<br>(p.R169Q) | homozygous | chr5:132384155 |
| M015 | Negative | inconclusive<br>(heterozygous<br>variant) | *BCKDHA* | MSUD type<br>1A (AR) | c.1312T>A<br>(p.Y438N) | heterozygous | chr19:41424582 |
| M016 | Positive | positive | *ACADM* | MCAD<br>deficiency<br>(AR) | c.985A>G<br>(p.K329E) | homozygous | chr1:75761161 |
| M017 | Positive | positive | *ACADM* | MCAD<br>deficiency<br>(AR) | c.985A>G<br>(p.K329E);<br>c.199T>C<br>(p.Y67H) | heterozygous<br>for both variants | chr1:75761161;<br>chr1:75732724 |

Seven participants previously diagnosed with phenylketonuria (PKU) by newborn screening had pathogenic variants in *PAH* (MIM: 612349); six of them were likely compound heterozygous for two different variants (ranging from missense, canonical splice-site, nonsense, and/or deletion variants). In view of their previously established diagnosis of PKU, we did not routinely perform parental testing to confirm the phase of these *PAH* variants. The seventh participant was likely homozygous for a pathogenic missense variant in *PAH* (c.194T>C (p.I65T)), although we did not rule out the possibility of a deletion *in trans*.

Seven participants previously diagnosed with medium-chain acyl-coA dehydrogenase (MCAD) deficiency (MIM: 201450) by newborn screening had pathogenic variants in *ACADM* (MIM 607008); five of these seven were homozygous for a well-known pathogenic missense variant (c.985A>G (p.K329E)) reported previously in individuals with MCAD deficiency^22–25^. Two of these seven were likely compound heterozygous for two different variants in the *ACADM* gene: c.985A>G (p.K329E) and c.799G>A (p.G267R); c.985A>G (p.K329E) and c.199T>C (p.Y67H).

One participant who was previously diagnosed with carnitine deficiency (MIM: 212140) by newborn screening was homozygous for a pathogenic *SLC22A5* (MIM: 603377) missense variant (c.506G>A (p.R169Q)) consistent with his clinical diagnosis^26–29^.

NGS-NBS analysis was deemed “negative” in 2 of 17 participants (12%) in the metabolic cohort. However, after unblinding, diagnostic analysis revealed suggestive, but inconclusive findings (Tables 1 and 2). One participant previously diagnosed with maple syrup urine disease (MSUD, MIM: 248600) was heterozygous for a pathogenic missense variant in the *BCKDHA* (MIM: 608348) gene (c.1312T>A (p.Y438N)) associated with classic, autosomal recessive MSUD^30–34^. Since we did not identify any additional variants in *BCKDHA* that might explain a genetic etiology for this participant’s disease, we classified this result as inconclusive. Another participant previously diagnosed with malonyl-CoA decarboxylase deficiency (MIM: 248360) was homozygous for a missense variant (c.1013T>C (p.L338P)) in *MLYCD* (MIM: 606761), the gene that encodes the malonyl-CoA decarboxylase enzyme, but this variant was classified as a VUS due to insufficient evidence. Of note, this infant was born prematurely at 23 weeks gestation, and required several repeat newborn screens due to inconclusive findings prior to eventual confirmation of malonyl-CoA decarboxylase deficiency at 7 weeks of age.

### Hearing loss cohort

NGS-NBS analysis was deemed to represent an abnormal, “positive screen” result in 5 of 28 participants (18%) in the hearing loss cohort (Tables 1 and 3). These individuals had a combination of pathogenic and/or likely pathogenic variants in genes implicated in hearing loss that, when blinded to cohort, were determined to be reportable in a screening setting. Two participants were each presumed to be compound heterozygous for two variants in the *USH2A* (MIM: 608400) gene based on ES data (c.1256G>T (p.C419F) and c.3686T>G (p.L1229Ter); c.4338_4339 delCT (p.C1447fs) and c.2299delG (p.E767fs)). In both cases, we subsequently confirmed that the variants were *in trans* by parental testing. One participant was homozygous for a 1-bp frameshift deletion (c.35delG (p.G12fs)) in *GJB2* (MIM: 121011; encoding connexin 26), a common pathogenic variant associated with DFNB1 nonsyndromic deafness (MIM: 220290)^35–38^. One participant was compound heterozygous for 2 pathogenic missense variants (c.626G>T (p.G209V) and c.1151A>G (p.E384G)) in *SLC26A4* (MIM: 605646), a gene associated with Pendred syndrome (MIM: 274600), an autosomal recessive form of syndromic hearing loss^39–43^. Finally, one participant was heterozygous for a pathogenic variant (c.5597C>T (p.T1866M)) in *TECTA* (MIM: 602574), a gene associated with autosomal dominant hearing loss (MIM: 601543)^44–46^. Of note, this infant passed her newborn hearing screen, but presented at one year of age with unilateral hearing loss that progressed to both ears. She was included in the study after referral by her audiologist, despite not fitting the original enrollment criteria, as her hearing loss was not detected by newborn screening.

**Table 3:**
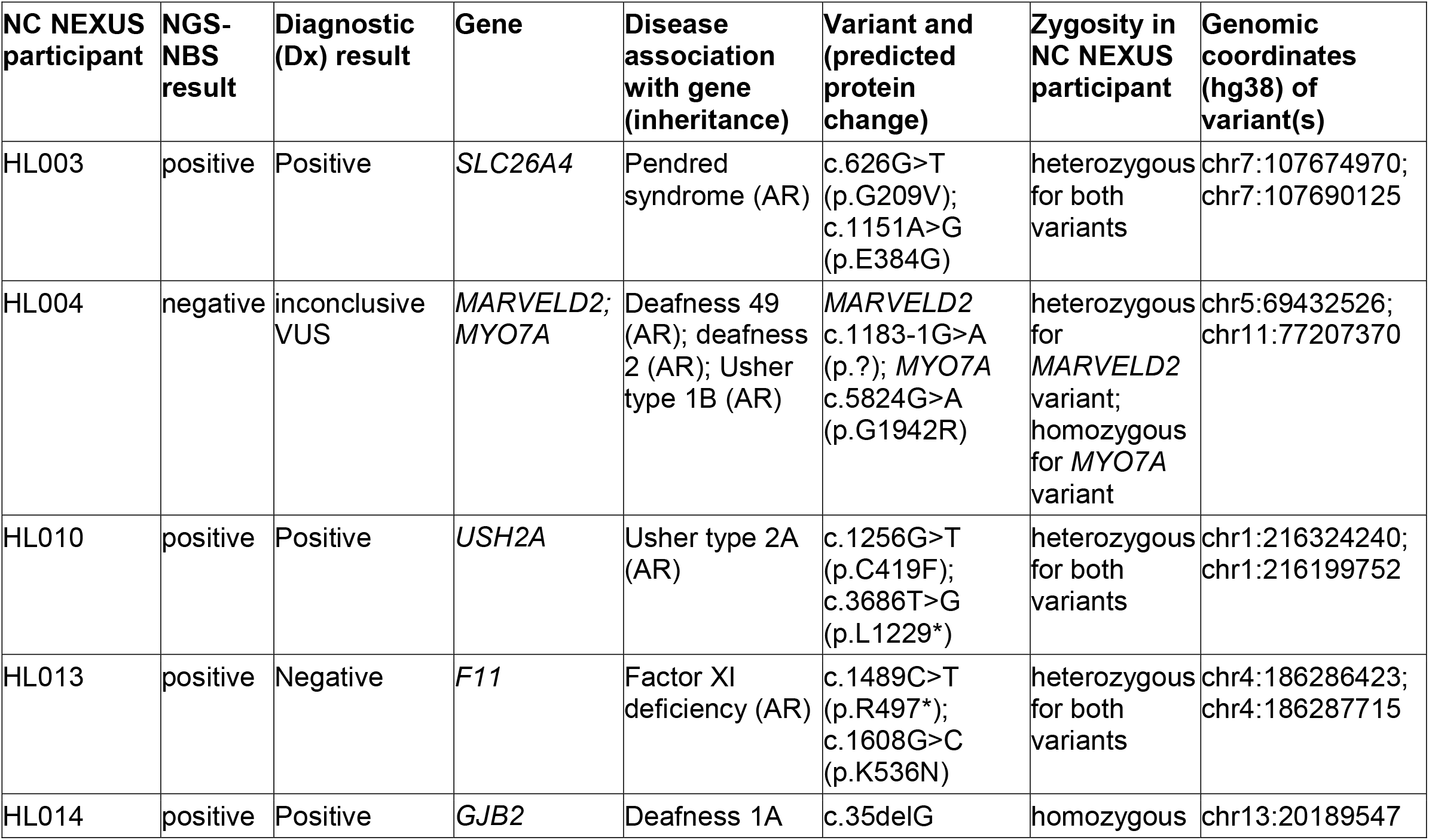
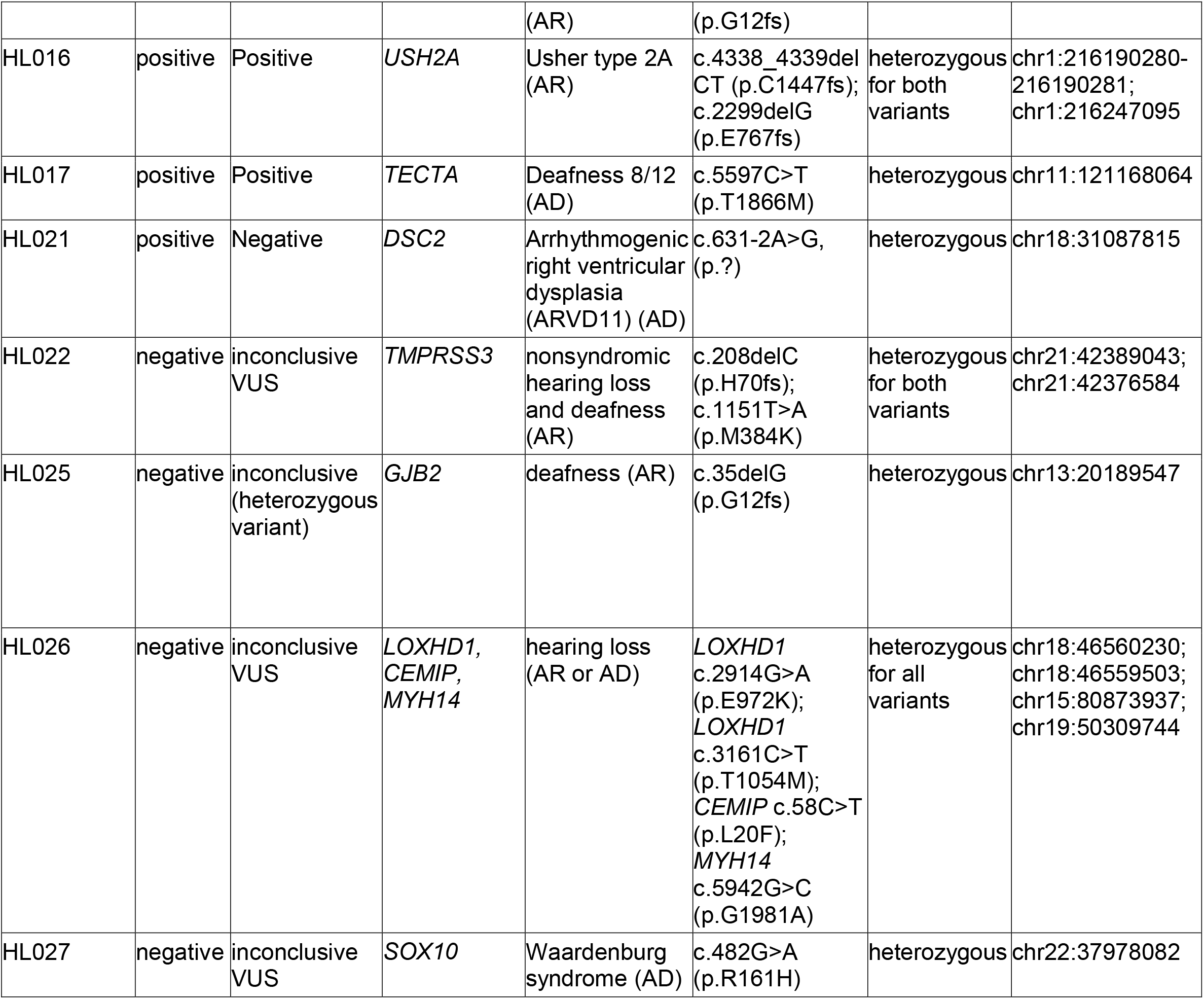
Participants in the NC NEUXS hearing loss cohort with positive or inconclusive findings. The NC NEXUS participant column includes “HL” (indicating hearing loss cohort) followed by the participant number. Each row in the table represents one NC NEXUS participant. Positive NGS-NBS results indicate a likely pathogenic or pathogenic variant found in the newborn screen. A positive diagnostic result indicates a likely pathogenic or pathogenic variant in a gene on the hearing loss list. An inconclusive diagnostic result is indicated by any variant of uncertain significance (VUS) finding or a heterozygous variant found in a gene associated with an autosomal recessive form of hearing loss. NGS-NBS= next-generation sequencing newborn screen; AR= autosomal recessive pattern of inheritance; AD= autosomal dominant pattern of inheritance.

In 5 of 28 participants in the hearing loss cohort, NGS-NBS screening analysis was deemed “negative.” However, after unblinding, diagnostic analysis identified 5 “inconclusive” results that may provide a possible explanation for their hearing loss (Tables 1 and 3). One participant was heterozygous for the pathogenic c.35delG (p.G12fs) deletion in *GJB2*. No additional pathogenic variants were detected in *GJB2*, and analysis in the CLIA-certified laboratory for intragenic *GJB6* (MIM: 604418) deletions was negative; however, we cannot rule out the presence of a second pathogenic variant in *GJB2* not detected by ES, such as a partial gene deletion or cryptic splice alteration caused by a non-coding variant.

A few other “inconclusive” findings were reported that did not provide definitive diagnostic results. One participant was heterozygous for four different missense VUSs in genes associated with hearing loss: two variants in *LOXHD1* (MIM: 613072) (c.2914G>A (p.E972K) and c.3161C>T (p.T1054M)); one variant in *CEMIP*, also known as *KIAA1199* (MIM: 608366) (c.58C>T (p.L20F)) and one variant in *MYH14* (MIM: 608568) (c.5942G>C (p.G1981A)). One participant was heterozygous for a missense VUS in *SOX10* (MIM: 602229) (c.482G>A (p.R161H)); a gene associated with autosomal dominant Waardenburg syndrome (MIM: 611584, 613266)^47^. One participant was homozygous for a missense VUS in *MYO7A* (MIM: 276903) (c.5824G>A (p.G1942R)); a gene associated with autosomal recessive syndromic and non-syndromic hearing loss (MIM: 276900, 600060)^48,49^. One participant was compound heterozygous for one likely pathogenic variant and one VUS in *TMPRSS3* (MIM: 605511): c.208delC (p.H70fs) and c.1151T>A (p.M384K); parental testing indicated that each variant was *in trans*.

We did not identify any P or LP variants on the diagnostic gene lists for 18 out of 28 (64%) participants in the hearing loss cohort; therefore, diagnostic analyses were deemed negative for identifying a genetic cause of hearing loss in these individuals (Table 1). Interestingly, one participant referred by audiologists to the hearing loss cohort had a rare syndromic diagnosis, Warsaw breakage syndrome (MIM: 613398), which is characterized by severe microcephaly and intellectual disability, growth restriction and sensorineural hearing loss due to cochlear hypoplasia^50,51^. Clinical quad exome sequencing of the similarly affected sibling, this participant and parents revealed a homozygous pathogenic variant in exon 13 of the *DDX11* (MIM: 601150) gene (c.1403dupT (p.S469fs)) in the participant and his sibling. This *DDX11* c.1403dupT (p.S469fs) variant was present in the NC NEXUS research ES data but was not examined because the *DDX11* gene was not included in either the NGS-NBS list or the hearing loss diagnostic list. If additional phenotypic information had been available at the time of molecular analysis, it is likely this variant would have been recognized as having diagnostic significance.

### NGS-NBS conditions

An abnormal, “positive” NGS-NBS result was also defined as a finding that predicted a childhood-onset actionable condition in any individual in the well-child cohort, or a condition unrelated to the indicated diagnosis of a member of one of the affected cohorts. NGS-NBS analysis was deemed “positive” in 4 of 106 participants (Table 4). One participant in the well-child cohort was heterozygous for a missense *LDLR* (MIM: 606945) variant (c.502G>A (p.D168N)) previously reported in autosomal dominant familial hypercholesterolemia (MIM: 143890)^52–58^. After disclosure of this finding, the parents stated that they were aware of a family history of hypercholesterolemia.

**Table 4:** Other actionable findings in the NC NEXUS NGS-NBS. The NC NEXUS participant column includes “M” (indicating inborn errors of metabolism cohort), “HL” (indicating hearing loss cohort) or “NB” (indicating well-child cohort) followed by the participant number. Each row in the table represents one NC NEXUS participant. Other actionable findings are indicated by variants found in the NGS-NBS that are associated with a disorder not previously known in the NC NEXUS participant. NGS-NBS= next-generation sequencing newborn screen; XL = X-linked inheritance pattern; AR= autosomal recessive pattern of inheritance; AD= autosomal dominant pattern of inheritance.

| NC NEXUS participant | NGS-NBS result | Gene | Disease association with gene (inheritance) | Variant and (predicted protein change) | Zygosity in NC NEXUS participant | Genomic coordinates (hg38) of variant(s) |
| --- | --- | --- | --- | --- | --- | --- |
| M003 | positive | *OTC* | OTC deficiency (XL) | c.1061T>G (p.F354C) | heterozygous | chrX:38421078 |
| NB012 | positive | *LDLR* | Familial hypercholesterolemia (AD or AR) | c.502G>A (p.D168N) | heterozygous | chr19:11105408 |
| HL013 | positive | *F11* | Factor XI deficiency (AR) | c.1489C>T (p.R497*), c.1608G>C (p.K536N) | heterozygous for both variants | chr4:186286423; chr4:186287715 |
| HL021 | positive | *DSC2* | Arrhythmogenic right ventricular dysplasia (ARVD11) (AD) | c.631-2A>G (p.?) | heterozygous | chr18:31087815 |

One female participant in the metabolic cohort, diagnosed with PKU, was a carrier of a missense variant (c.1061T>G (p.F354C)) in the ornithine transcarbamylase (*OTC*, MIM: 300461) gene. This variant was previously reported in mild OTC deficiency (MIM: 311250)^59,60^, an X-linked disease with variable expressivity in carrier females. Although this participant was diagnosed with PKU by the standard newborn screen at birth, the OTC deficiency was an unexpected finding arising from the ES analysis. Her ammonia levels are normal, and the variant was also found in her unaffected grandfather. Her younger male sibling was prenatally diagnosed with the variant, which enabled early measurement of ammonia level immediately after delivery and appropriate ongoing health supervision. So far, he has been asymptomatic. One hearing loss cohort participant was heterozygous for a canonical splice-site variant in the *DSC2* (MIM: 125645) gene (c.631-2A>G (p.?)). This variant has been reported previously in individuals with autosomal dominant arrhythmogenic right ventricular dysplasia (MIM: 610476)^61^. This child has had a normal echo and it was recommended that the child be followed by a cardiologist. Another hearing loss cohort participant had two variants in the *F11* (MIM: 264900) gene (nonsense variant c.1489C>T (p.R497Ter) and missense variant c.1608G>C (p.K536N)). Both variants have been reported previously in individuals with autosomal recessive factor XI deficiency (MIM: 612416)^62–64^. This child has a history of frequent episodes of epistaxis (nosebleeds), one requiring cauterization. It was recommended that she be evaluated by a hematologist, and it was also recommended that precautions are taken prior to any surgical procedures.

### Parental decision-making and additional genomic information

A total of 65 participants were randomized into the experimental decision arm of the study, viewed the decision aid, and attended the study visit with a genetic counselor. Over 90% of these parents chose to learn at least one category of additional genomic information: 47 (72.3%) requested all three categories of additional information, 7 requested additional findings for both adult-onset actionable conditions and carrier status, 2 requested additional findings for the adult-onset actionable category only, 1 requested carrier status only, 1 requested findings for the adult-onset actionable and childhood-onset nonactionable conditions categories, and 1 requested the childhood-onset nonactionable category only. The parents of 6 participants (9.2%) elected not to receive any of the additional information categories.

Two participants had additional findings in the adult-onset actionable category (Supplemental Table 2). A participant in the metabolic cohort was heterozygous for a pathogenic deletion *RAD51C* (MIM: 602774) variant (c.905-2_905-1delAG (p.?)), which is predicted to alter a splice site and has been reported previously in autosomal dominant familial ovarian cancer (MIM: 613399)^65,66^. This participant had a known family history of breast cancer, but no known family history of ovarian cancer. A participant in the well-child cohort was heterozygous for a nonsense c.7480C>T (p.R2494Ter) variant in *BRCA2* (MIM: 600185) that is a known pathogenic variant associated with autosomal dominant familial breast and ovarian cancer (MIM: 612555)^67–74^. This participant’s parents reported a maternal family history of breast and pancreatic cancer. The child’s mother was referred to adult genetics.

One participant in the hearing loss cohort had an additional finding for the childhood-onset nonactionable conditions category (Supplemental Table 2). This participant was hemizygous for a likely pathogenic nonsense variant (c.741G>A (p.W247Ter)) in the *OCRL* (MIM: 300535) gene. This gene is associated with Lowe syndrome^75^ (MIM: 309000), and further correlation with clinical findings indicated that the participant has multiple features consistent with Lowe syndrome. Indeed, we subsequently learned that the patient had been seen in clinical genetics and clinical exome sequencing was considered, but not performed due to concerns about cost. Although hearing loss is not a prominent feature of Lowe syndrome, there is at least one previous case report^76^ involving an individual with Lowe syndrome who also had hearing loss, suggesting that the finding may provide a comprehensive diagnostic result for this participant’s symptoms. The mother of this patient is a carrier of the variant and cascade testing of other family members has been negative.

The number of carrier findings in children whose parents requested this category ranged from 0-7 variants, with an average of 1.8 reportable carrier findings per individual (Figure 1, Supplemental Table 2). A total of 100 out of 109 (92%) potential carrier findings detected by ES were confirmed by orthogonal testing in a CLIA-certified laboratory and reported. The nine variants that failed to be confirmed by orthogonal testing had low read depth for both alleles in the ES data. We observed several recurrent carrier findings; the most common were variants in the genes *HFE* (MIM: 613609), *GALT* (MIM:606999), *SERPINA1* (MIM:107400), and *RBM8A* (MIM: 605313) (Figure 2). A total of 11 participants carried the *HFE* c.187C>G (p.H63D) variant, while 4 carried the *HFE* c.845G>A (p.C282Y) variant, both associated with hereditary hemochromatosis (MIM: 235200)^77,78^. Four participants each carried the *SERPINA1* S or Z alleles (c.863A>T (p.E288V) and c.1096G>A (p.E366K)), respectively) associated with alpha-1-antitrypsin deficiency (MIM: 613490)^79^. Five participants were carriers for variants in *GALT* that are associated with the Duarte 2 allele^80–82^ (including 1 missense variant and 3 intronic variants), while 2 participants were carriers for a variant associated with classic galactosemia (MIM: 230400), c.563A>G (p.Q188R)^83,84^. Five participants carried a hypomorphic variant located in the 5□ untranslated region (UTR) in *RBM8A* (c.-21G>A (p.?)), associated with thrombocytopenia-absent radius (TAR) syndrome (MIM: 274000)^85,86^. Additionally, we identified three carriers of pathogenic *CFTR* (MIM: 602421) variants associated with cystic fibrosis^87–90^ (MIM: 219700); two participants carried the *CFTR* c.1521_1523delCTT (p.F508del) founder variant and one participant carried the *CFTR* c.1519_1521delATC (p.I507del) variant (Supplemental Table 2).

**Figure 1.**
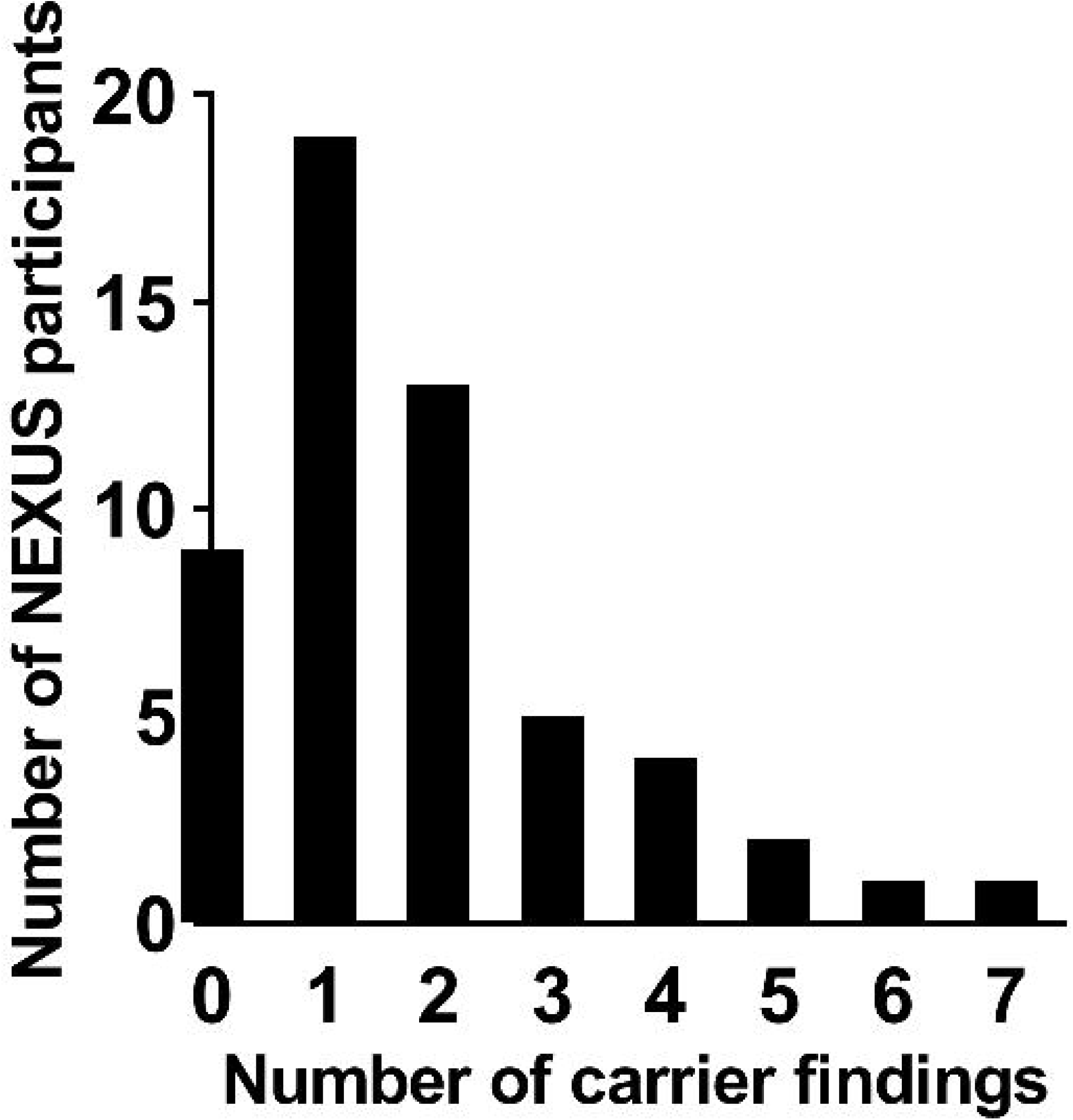
Number of carrier findings for NC NEXUS participants that were randomized into the experimental group and elected to receive carrier findings. Number of carrier findings is represented on the x-axis and the number of NC NEXUS participants is represented on the y-axis.

**Figure 2.**
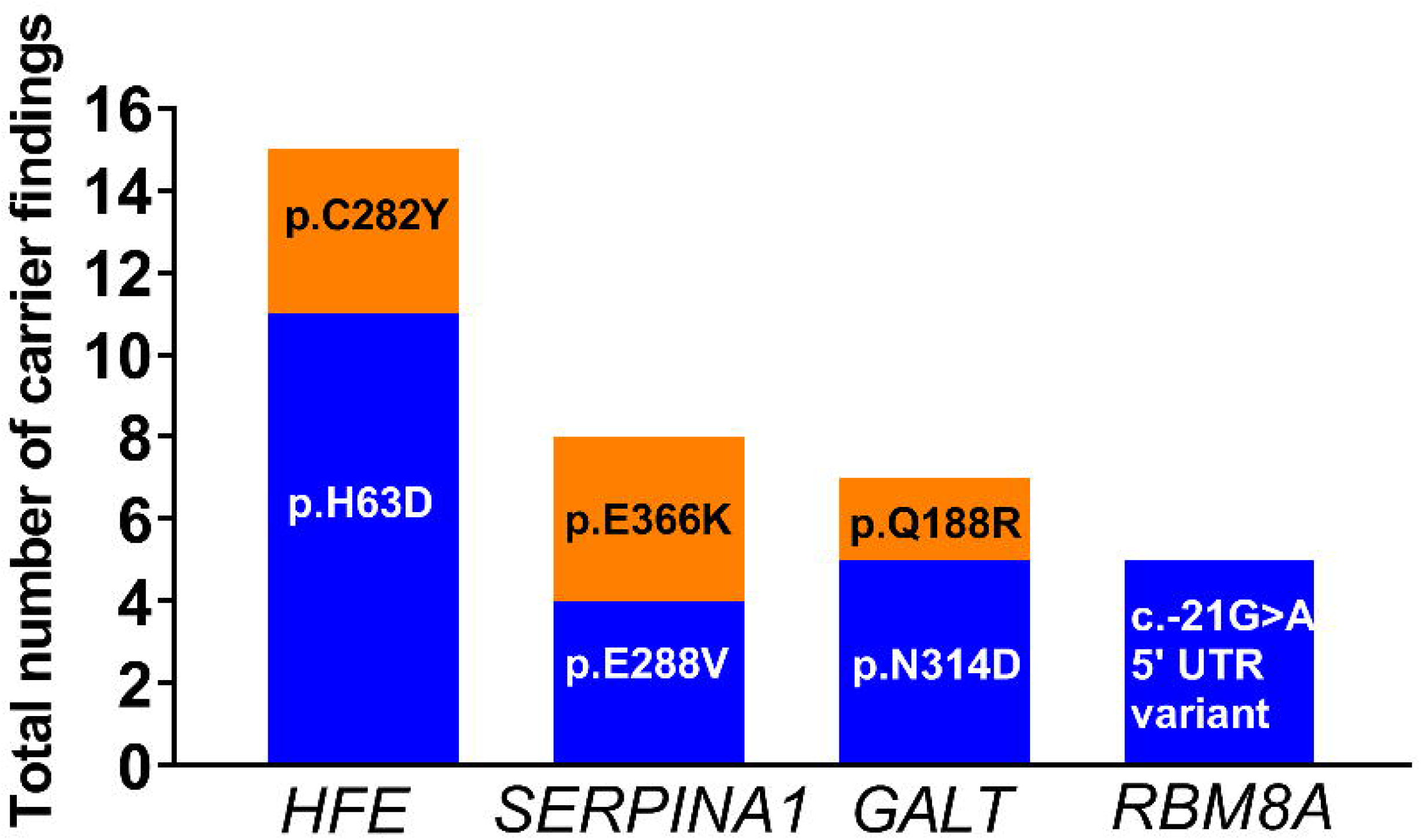
Variants in *HFE, SERPINA1, GALT* and *RBM8A* were the most frequently observed carrier findings. Variants/predicted protein changes and genes in which they are located are represented on the x-axis. Total number of carrier findings in the NC NEXUS study is represented on the y-axis.

## Discussion

The potential use of genome-scale sequencing to screen healthy individuals for monogenic disorders--before symptoms manifest--promises the tantalizing possibility of ameliorating these conditions, but also raises numerous challenges. Among the barriers to the use of genomic sequencing in any screening endeavor is the performance of this technology and our ability to interpret variants in the absence of phenotypic indications. Other critical issues include the kinds of genomic information that should be offered, how parental informed consent can be obtained, and how the overall process and the potential results will impact families. To address these questions, the NC NEXUS study enrolled a cohort of 106 children: 17 with inborn errors of metabolism, 28 with hearing loss, and 61 healthy newborns. We explored the potential use of ES in the context of newborn screening, both from the standpoint of analytic performance as well as parental preferences for genomic information.

### Analytic Performance

In order to assess how well ES would perform for predictive purposes, molecular analysts were initially blinded to the cohort status of the individual, enabling us to simulate the conditions of a true screening test, in which the individual is assumed to be at population risk. We did not perform trio analyses, which can be helpful for identifying *de novo* and compound heterozygous variants, although we did conduct follow-up parental sequencing whenever possible where compound heterozygosity was suspected. From the standpoint of implementation workflows, it is reasonable to attempt to obtain trio samples in order to maximize the diagnostic yield of clinical diagnostic sequencing; however, comprehensive trio sample collection would be impractical within current NBS practices.

Screening is unlike indication-based diagnostic testing, in which the analyst can utilize phenotypic information to judge which variants are most likely to be relevant, and where the disclosure of inconclusive results (variants of uncertain significance, single heterozygous variants in genes associated with recessive conditions, etc.) can still be informative and allow for more detailed clinical follow-up. However, in a population at low risk for any given monogenic disorder, disclosure of genomic findings with lower certainty will inevitably lead to an extremely high false positive rate, and we therefore set relatively stringent thresholds for the results to qualify as a “positive screen.” This decision reflects the inevitable balance between increasing sensitivity to maximize case finding, versus establishing stringent thresholds to reduce false positives. As expected, a small number of participants (4 out of the total 106) had positive NGS-NBS screening results (beyond those that would be expected for those in the affected cohorts), but the results have implications for their health supervision. These findings exemplify the expansion of the possible range of conditions that could be identified in newborns, and thereby influence future health care and screening to ultimately improve clinical outcomes. These findings had implications for other family members, but longitudinal follow-up of a larger number of screened individuals will be required to assess outcomes, clinical utility, and the economic value of NGS-NBS. Among the affected cohorts, the screening analysis predicted the presence of inborn errors of metabolism in 15 out of 17 individuals and hearing loss in 5 out of 28 individuals. Thus, at a population level, it is unlikely that genomic sequencing could replace current screening based on biochemical measurements or audiology testing.

In addition to a purely screening mode of analysis, genomic information could also be used in conjunction with phenotypic information (such as a metabolic screening assay or hearing screen) to perform a “secondary” or “indication-based” analysis. Therefore, after the analyst determined the “screening” results for a given individual, they were unblinded to the cohort that the individual was from, and a more sensitive diagnostic analysis was conducted. The yield of sequencing improved somewhat in the context of indication-based analysis: the two remaining individuals in the metabolic cohort and several members of the hearing loss cohort had highly suggestive, but “inconclusive” results (e.g. a single heterozygous pathogenic variant, a homozygous missense VUS in a gene associated with an autosomal recessive disease). These findings suggest that sequencing might be useful as an adjunct to traditional NBS methods, and that with improved detection of variants (such as non-canonical splicing variants, copy number variants or rearrangements) and more extensive interpretive databases, the positive predictive value of genomic screening may improve.

A potential explanation for the relatively low diagnostic yield of ES for participants in the hearing loss cohort is that numerous factors^91–95^ contribute to hearing loss, including non-genetic factors (i.e. environmental, infections, premature birth, etc.) that are estimated to account for approximately 20% of cases of prelingual hearing loss^96^. The diagnostic rate in our study likely reflects the genetic heterogeneity of the monogenic forms of hearing loss^97,98^ as well as the incomplete knowledge of pathogenic variants. Ascertainment bias also may have led to a larger fraction of the hearing loss cohort having complex syndromic conditions that led the referring providers to consider exome sequencing as a useful diagnostic test. Other hearing loss patients may already have had panel testing, thus depleting the enrolled study population of the more commonly identified nonsyndromic hearing loss conditions. In addition, certain genetic forms of hearing loss are caused by copy number variants or mitochondrial DNA variants^96,99^, which were not assessed in this study. Due to the heterogeneity of the etiologies of hearing loss, it seems clear that phenotypic screening at birth will remain necessary, even if detection of the genetic mechanisms improves.

On the other hand, many types of hearing loss that would be detectable with genomic sequencing (for example, hearing loss due to pathogenic or likely pathogenic variants in *TECTA* as shown in one NC NEXUS patient) do not have congenital onset and would therefore be missed at birth. The pathogenic *TECTA* variant finding demonstrates the potential for NGS-NBS to detect hearing loss risk in children who are not identified through traditional newborn hearing screening. The identification of a genetic risk for hearing loss in this setting could be followed up with more aggressive audiology evaluations during infancy and early childhood in order to facilitate interventions and reduce the chance of language impairment. In other cases, hearing loss is associated with a broader syndromic condition, for example the NC NEXUS patients with Warsaw Breakage Syndrome or Lowe Syndrome. These examples raise intriguing points regarding the implementation of genomic screening in newborns, where testing in the context of a specific phenotype can be a powerful diagnostic tool enabling families and clinicians to better manage an individual’s health condition. However, it is unclear whether routine screening of all healthy newborns for such conditions would be useful in a broader context without clinical interventions to ameliorate symptoms; in such cases, parental input will be critical.

### Parental decision-making and additional genomic findings

One question raised by the prospect of genomic sequencing in newborns is how parents will be engaged in the process of informed decision making. After using an online decision aid and discussion with a genetic counselor, over 72% of parents in our study who were eligible to learn additional findings chose to learn information in all three categories. The carrier findings reported in this study were similar to other studies^10,100–103^. Interestingly, many of the variants we observed are milder or hypomorphic variants; in order to cause disease, these alleles must be *in trans* with a more severe variant in the same gene (e.g. the *RBM8A* c.-21G>A (p.?) variant and the *HFE* c.187C>G (p.H63D) variant). These hypomorphic variants complicate the analysis and the reporting of carrier findings. In addition, identifying carrier status in a newborn is not fully informative with regards to carrier status in the parents or siblings without further analysis, which decreases the clinical utility of reporting carrier findings in infants. Parental preferences identified in this study may have limited generalizability given that ∼40% of the participants already had health conditions with a possible genetic etiology. Other factors may also have influenced whether parents decided to enroll their child to participate in the NC NEXUS study (e.g. parents’ education level, parents’ general feelings about research studies). Much information remains to be learned about parental decisions in diverse contexts.

### Conclusions and future directions

Clinical applications of genomic sequencing technologies offer great opportunities in both diagnostic testing and screening. In the context of NBS, sequencing-based approaches cannot fully replace biochemical or phenotypic screens because of etiologic heterogeneity and the challenges of variant interpretation. However, sequencing approaches have the advantage of being able to identify virtually any condition with a known genetic cause. Thus, augmentation of newborn screening with some form of genomic sequencing seems inevitable. In the near term, sequencing of genes already associated with conditions that are screened for in traditional NBS may help to improve clinical sensitivity and specificity. Expanding the current NBS panel to include other actionable conditions detectable only by sequencing could further enhance the public health benefits of NBS, if interpretive challenges can be overcome in order to balance case detection versus false positives. Providing additional genomic information beyond the most actionable conditions, while potentially of interest to many parents, may increase the complexity of informed consent and thereby serve to distract from the primary health benefits of NGS-NBS. There will be significant costs, ethical considerations and implementation challenges involved in conducting genome-scale sequencing in healthy newborns. These challenges may limit broad application across the entire population, thus failing to deliver on the promise of the Human Genome Project. We should therefore continue to explore innovative approaches to NGS-NBS that will enable the most comprehensive adoption in the general population, improve outcomes to the greatest extent possible, and maximize societal benefits in a cost-effective manner.

## Data Availability

Variants reported in this manuscript have been submitted to the ClinVar database.

http://ncbi.nlm.nih.gov/clinvar/

## Description of Supplemental Data

Supplemental data include 2 tables.

## Acknowledgments

We thank the families who participated in the study. This work was funded by the National Human Genome Research Institute and the *Eunice Kennedy Shriver* National Institute of Child Health and Human Development of the National Institutes of Health as part of the Newborn Sequencing in Genomic Medicine and Public Health (NSIGHT) Program, U19 HD077632. We gratefully acknowledge the UNC BioSpecimen Processing Facility and the UNC High-Throughput Sequencing Facility for their work in DNA extraction, sample preparation and exome sequencing, and Manyu Li and the UNC Hospitals Clinical Molecular Genetics Laboratory. We also thank Jason Reilly, Ian Wilhelmsen, Kirk Wilhelmsen, Dylan Young and the Renaissance Computing Institute (RENCI) at UNC for bioinformatics analysis and expertise; Alexandra Arreola and Claire Edgerly for contributions to molecular analysis; and Lonna Mollison and Kathleen Wallace for their contributions to curating the NGS-NBS and additional findings gene lists.

ClinicalTrials.gov Identifier: NCT02826694

## Web Resources

Online Mendelian Inheritance in Man: Omim.org

ClinVar database: ncbi.nlm.nih.gov/clinvar/

UCSC genome browser: genome.ucsc.edu

Genome Aggregation Database: gnomad.broadinstitute.org

Exome Aggregation Consortium Browser: exac.broadinstitute.org

ClinGen Variant Pathogenicity Curation Interface: curation.clinicalgenome.org

